# Risk factors for severe disease in patients admitted with COVID-19 to a hospital in London, England: a retrospective cohort study

**DOI:** 10.1101/2020.09.24.20200337

**Authors:** J. W. Goodall, T. A. N. Reed, M. Ardissino, P. Bassett, A. M. Whittington, D. L. Cohen, N. Vaid

## Abstract

COVID-19 has caused a major global pandemic and necessitated unprecedented public health restrictions in almost every country. Understanding risk factors for severe disease in hospitalized patients is critical as the pandemic progresses.

This observational cohort study aimed to characterize the independent associations between the clinical outcomes of hospitalized patients and their demographics, comorbidities, blood tests and bedside observations. All patients admitted to Northwick Park Hospital, London, United Kingdom between 12 March and 15 April 2020 with COVID-19 were retrospectively identified. The primary outcome was death. Associations were explored using Cox proportional hazards modelling.

The study included 981 patients. The mortality rate was 36.0%. Age (adjusted hazard ratio (aHR) 1.53), respiratory disease (aHR 1.37), immunosuppression (aHR 2.23), respiratory rate (aHR 1.28), hypoxia (aHR 1.36), Glasgow Coma Score <15 (aHR 1.92), urea (aHR 2.67), alkaline phosphatase (aHR 2.53), C-reactive protein (aHR 1.15), lactate (aHR 2.67), platelet count (aHR 0.77) and infiltrates on chest radiograph (aHR 1.89) were all associated with mortality.

These important data will aid clinical risk stratification and provide direction for further research.

## Introduction

COVID-19, the illness caused by infection with severe acute respiratory syndrome coronavirus 2 (SARS-CoV-2), has caused a global pandemic and has placed an unprecedented strain on healthcare systems worldwide. Early studies from China found that older age, male sex and a number of comorbidities (notably hypertension, cardiovascular disease and respiratory disease) [1–6], were associated with poorer outcomes. Furthermore, a number of blood test abnormalities such as raised inflammatory markers [3,7–11] and lymphopenia [10,12] have been associated with worse outcomes.

In the UK, two large cohort studies have corroborated the associations between older age, male sex and pre-existing comorbidities with poorer outcomes [13,14]. However, these studies included demographic and morbidity characteristics only, with no data to indicate disease severity at the time of presentation to hospital. The inclusion of investigations and observations that indicate clinical condition at presentation allows for increased acuity of predictive models. The aim of this large UK inpatient cohort study is to explore the association between results of admission laboratory tests and clinical observations, alongside demographic and morbidity characteristics, with clinical outcomes of patients hospitalized with COVID-19. All patients admitted with COVID-19 in the time period were enrolled thus avoiding the selection bias seen in other cohorts.

## Methods

### Study Population

Northwick Park Hospital is a 658 bedded district general hospital serving an ethnically and socioeconomically diverse population in North West London [15]. This observational cohort study retrospectively identified all adult patients (age ≥ 18 years) admitted to this hospital between 12 March and 15 April 2020 who were confirmed to be positive for SARS-CoV-2 by RT-PCR of pooled nasal and pharyngeal swabs. Patients were tested for COVID-19 in accordance with contemporaneous Public Health England guidelines [16]. Advice on which patients should be tested remained constant throughout the study period. Patients were admitted to hospital at the discretion of the admitting physician and managed using best supportive treatment, including high flow oxygen, antibiotics at the discretion of the treating physician, continuous positive airway pressure (CPAP), invasive ventilation and organ support as needed. This study had National Research Ethics approval (REC 20/NW/0218, IRAS 282630) and was carried out in accordance with the Declaration of Helsinki and the principles of Good Clinical Practice.

### Variables

Data were collected from electronic patient records and entered into a securely stored database. This was pseudonymized for data analysis. Age, sex and ethnicity were recorded. Ethnicity was categorized as White, Black, Asian or Other (including mixed ethnicity). Comorbidities and medications which previous research had suggested might be associated with prognosis were extracted, including: hypertension, heart failure (HF), ischaemic heart disease (IHD), diabetes mellitus (DM), active malignancy and respiratory disease (including asthma, chronic obstructive pulmonary disease (COPD), pulmonary fibrosis, and obstructive sleep apnoea). Use of angiotensin converting enzyme inhibitors (ACEi), angiotensin-receptor blockers (ARB), metformin, and immunosuppressive medications (including for autoimmune disease and anti-rejection drugs for solid organ transplant) were recorded. Information on comorbidities from admission documentation was cross-referenced with recent inpatient and specialist communications.

The following observations were collected on arrival to hospital: heart rate, blood pressure, temperature, respiratory rate, Glasgow Coma Scale (GCS), and hypoxia. Hypoxia was defined as oxygen saturations of <94% on air or the need for supplemental oxygen to maintain saturations ≥ 94%. Exceptions were made for two patients with established COPD who were known to retain carbon dioxide and had pre-COVID-19 saturations <94%.

The following blood test results were collected: haemoglobin, platelet count, lymphocyte count, neutrophil count, potassium, sodium, creatinine, urea, alkaline phosphatase (ALP), alanine aminotransferase (ALT), bilirubin, albumin, C-reactive protein (CRP), and lactate. As lactate levels may change rapidly in response to clinical interventions lactate was only recorded if taken within four hours of admission; all other blood results represent the first recorded on admission.

The presence or absence of formally reported pulmonary infiltrates on plain chest radiographs taken on admission was recorded.

Medications potentially active against SARS-CoV-2 as part of research trials including remdesivir, dexamethasone, hydroxychloroquine, lopinavir-ritonavir, azithromycin and tocilizumab were recorded.

### Outcomes

Outcomes were ascertained for all patients on the census date of 13 May 2020, 28 days following the latest admission date. For all patients who were discharged from hospital alive, the end of follow-up date was considered to be the census date.

The primary outcome was death during the study period. Two secondary outcomes were considered. First, the composite of death or invasive mechanical ventilation (IMV). Second, the composite of death, IMV or CPAP.

### Statistical analysis

Descriptive statistical analyses were performed on all of the population characteristics. Kaplan-Meier methods were used to calculate and graph cumulative survival.

Analyses examining factors associated with survival times were performed using a Cox proportional hazards model. Variable selection was executed by a multi-stage procedure. First, independent variables were examined in a series of univariable analyses. Where necessary, higher-order terms were used to better describe the relationship between continuous variables. Continuous variables found to have a heavily positively skewed distribution were analysed on the log^10^ scale.

In the second stage, only factors found to show association with survival times were included. A backwards selection procedure was used to retain the significant variables in the final model.

To visually display the effect of the combinations of risk factors on outcomes, patients were allocated to three equal-sized groups based on their predicted risk from the multivariable regression model. These outcomes were displayed as tertiles on a Kaplan-Meier plot.

Forty-one patients did not have a positive swab result until >5 days after admission. We were unable to accurately establish the reason for this, but potential explanations include initial false-negative swabs and presentations that did not initially fit the contemporaneous case definition of COVID-19. To explore this further, sensitivity analyses were performed with these patients excluded.

Statistical analysis was performed using STATA v15.1 (StataCorpLLC, Texas, USA). STATA codes can be obtained from the authors. Data on all COVID-19 admissions to the Hospital have already been shared with Public Health England. Application for further patient level data should be addressed to the Hospital’s Research Committee.

## Results

### Demographics and characteristics

Nine-hundred-and-eighty-one patients were included in the study. The median age was 69 (IQR 56-80) years, 64.3% were male, and 776 (79.1%) had at least one documented comorbidity. The cohort’s full demographic and comorbidity data are shown in Table 1.

**Table 1:** Demographics, comorbidities and medication usage.

| Variable (no. of patients with data available if data missing) | All | Alive | Died |
| --- | --- | --- | --- |
| **Demographics** |  |  |  |
| Age, Median (IQR) | 69 (56-80) | 63 (51-75.5) | 78 (68-85) |
| Sex (981) |  |  |  |
| Female | 350 (35.7) | 226 (36.0) | 124 (35.0) |
| Male | 631 (64.3) | 401 (64.0) | 230 (65.0) |
| Ethnicity (824) |  |  |  |
| White | 250 (25.5) | 150 (23.9) | 100 (28.2) |
| Black | 136 (13.9) | 95 (15.2) | 41 (11.6) |
| Asian | 371 (37.8) | 241 (38.4) | 130 (36.7) |
| Other | 67 (6.8) | 41 (6.5) | 26 (7.3) |
| **Comorbidities and medication use** |  |  |  |
| Any comorbidity | 776 (79.1) | 459 (73.2) | 317 (89.5) |
| Diabetes | 376 (38.3) | 218 (34.8) | 158 (44.6) |
| Hypertension | 486 (49.6) | 275 (43.9) | 211 (59.6) |
| Heart failure | 97 (9.9) | 47 (7.5) | 50 (14.1) |
| Ischemic heart disease | 143 (14.6) | 74 (11.8) | 69 (19.5) |
| Active malignancy | 47 (4.8) | 24 (3.9) | 23 (6.6) |
| Respiratory Disease - Any | 204 (20.8) | 119 (19.0) | 85 (24.0) |
| Asthma | 80 (8.1) | 52 (8.3) | 28 (7.9) |
| COPD | 45 (4.6) | 24 (3.8) | 21 (5.9) |
| OSA | 17 (1.7) | 8 (1.3) | 9 (2.5) |
| Pulmonary fibrosis | 14 (1.4) | 4 (0.6) | 10 (2.8) |
| Other | 48 (4.9) | 31 (4.9) | 17 (4.8) |
| Other Comorbidity | 198 (20.2) | 101 (16.1) | 97 (27.4) |
| ACEi / ARB | 255 (26.0) | 151 (24.1) | 104 (29.4) |
| Metformin | 210 (21.4) | 136 (21.7) | 74 (20.9) |
| Immunosuppression | 31 (3.2) | 15 (2.4) | 16 (4.5) |

ACEi/ARB: angiotensin converting enzyme inhibitor/angiotensin II receptor blockers. COPD: Chronic obstructive pulmonary disease. IQR:
interquartile range OSA: Obstructive sleep apnoea
Values are presented as number (percentage) unless otherwise stated.

Thirty patients received trial drugs for COVID-19: one received azithromycin alone; one received azithromycin with tocilizumab; ten received dexamethasone; ten received hydroxychloroquine; four received lopinavir-ritonavir and four received remdesivir.

### Laboratory findings

A raised CRP was seen in 96.2%, and 60% had raised platelets, consistent with inflammation. A significant number of patients had hypernatraemia (58.0%), raised urea levels (40.7%), and lymphopenia (76%).

Almost all patients (97.9%) had a chest radiograph taken on admission and of these 75.0% were reported as having pulmonary infiltrates. (The blood results are available in the supplementary material).

### Cohort outcomes

Median follow-up was 37 days (range 10-46). By the censoring date, 354 (36.0%) had died. A Kaplan-Meier survival curve is shown in Figure 1. Of the 627 patients alive at the end of the study period, 566 (90.3%) had been discharged from hospital, 13 (2.1%) had been discharged to rehabilitation facilities and 48 (7.7%) remained on acute wards.

**Figure 1:**
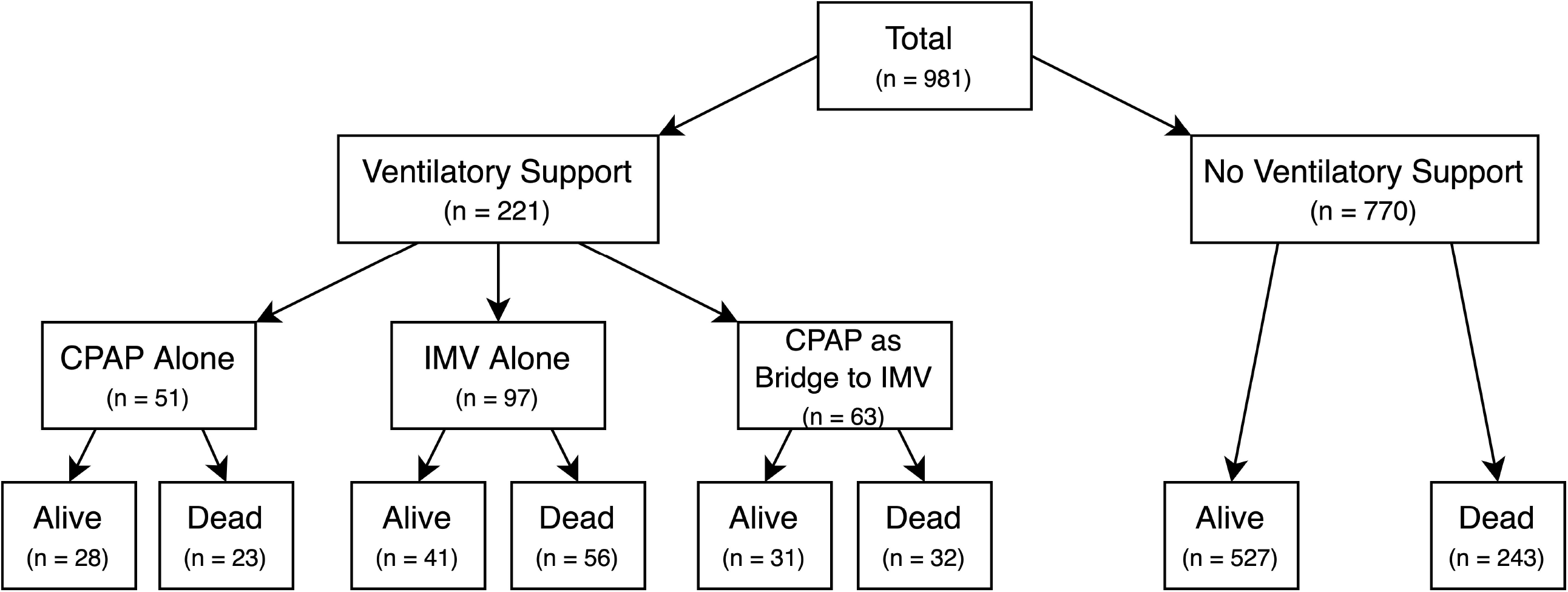
Kaplan-Meier plot of time to death (reduced y-axis scale). Blue shading corresponds to 95% confidence intervals

For the 579 discharged alive the median duration of stay was 8 days (range 0-46).

Out of 981 patients admitted in the study period, 114 (11.6%) required CPAP and 160 (16.3%) IMV. Notably, 28 patients (24.6%) who required CPAP survived without requiring IMV. The cohort outcomes are shown in Figure 2.

**Figure 2:**
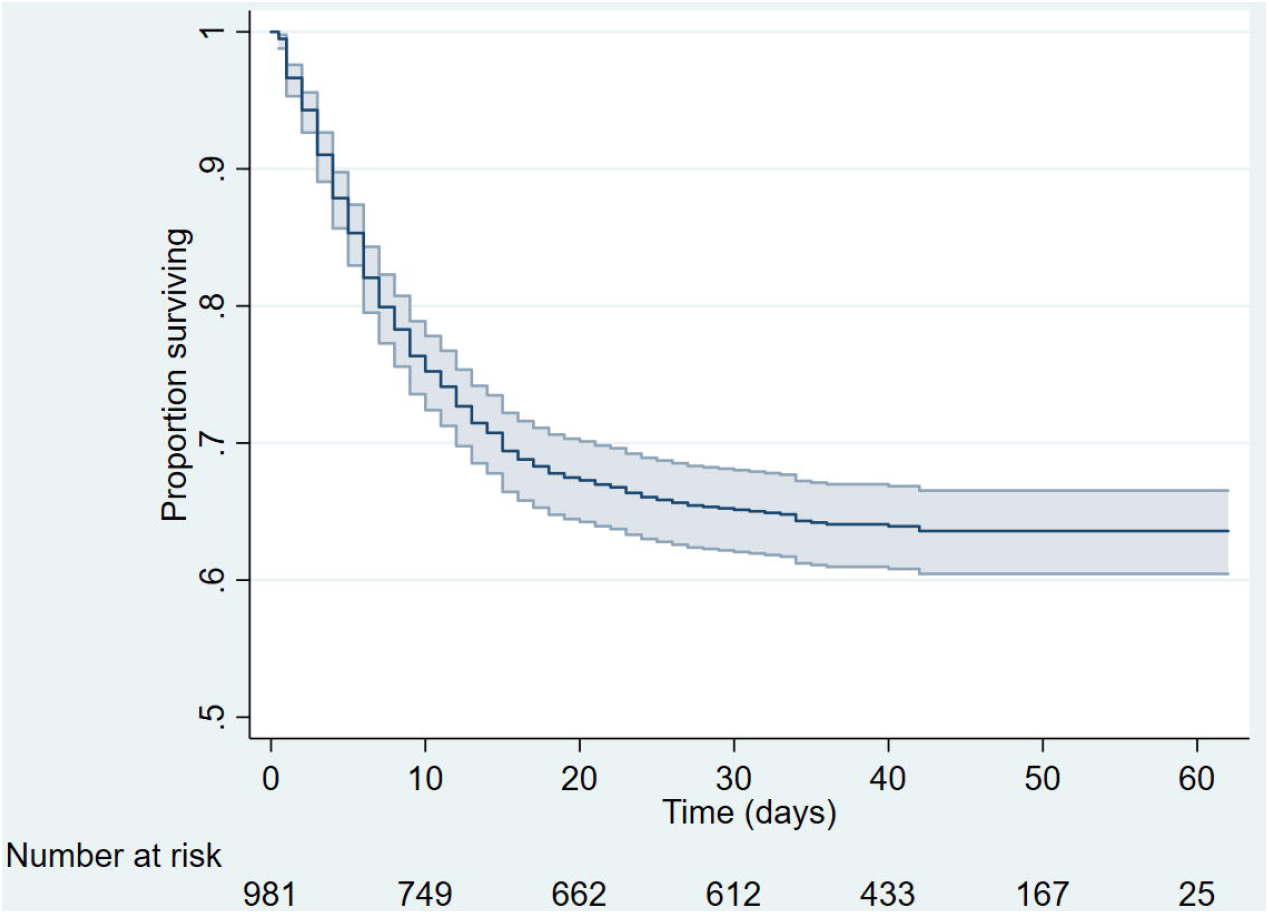
Cohort outcomes. CPAP: continuous positive airways pressure. IMV: invasive mechanical ventilation

### Primary outcome

The association between baseline demographic, clinical, laboratory, and radiological features with death, the primary outcome was examined. In multivariable analysis 12 variables retained statistical significance. The final model was based on data from 741 of the 981 patients, due to missing values for some variables. These results are shown in Table 2.

**Table 2:** primary outcome – time to death.

| Variable | Risk of death |  |  |  |
| --- | --- | --- | --- | --- |
|  | Univariable analysis |  | Multivariable analysis (n=772) |  |
|  | Hazard ratio<br>(95% CI) | p value | Adjusted hazard ratio<br>(95% CI) | p value |
| **Demographics** |  |  |  |  |
| Age <sup>(**)</sup> | 1.58 (1.46, 1.71) | <0.001 | 1.53 (1.37, 1.71) | <0.001 |
| Male sex (vs. female sex) | 1.02 (0.82, 1.23) | 0.88 |  |  |
| Ethnicity |  |  |  |  |
| White (reference) | 1 | 0.25 |  |  |
| Black | 0.71 (0.49, 1.02) |  |  |  |
| Asian | 0.84 (0.64, 1.08) |  |  |  |
| Other | 0.93 (0.61, 1.44) |  |  |  |
| **Comorbidities and Medication Usage** |  |  |  |  |
| Diabetes (vs. none) | 1.42 (1.15, 1.75) | 0.001 |  |  |
| Hypertension (vs. none) | 1.64 (1.33, 2.03) | <0.001 |  |  |
| Heart failure (vs. none) | 1.72 (1.27, 2.32) | <0.001 |  |  |
| Ischaemic heart disease (vs. none) | 1.61 (1.24, 2.09) | <0.001 |  |  |
| Active malignancy (vs. none) | 1.47 (0.97, 2.25) | 0.07 |  |  |
| Respiratory disease (vs. none) | 1.28 (1.00, 1.63) | 0.05 | 1.37 (1.03, 1.81) | 0.03 |
| ACEi / ARB (vs. none) | 1.25 (0.99, 1.57) | 0.06 |  |  |
| Metformin (vs. none) | 0.97 (0.75, 1.25) | 0.81 |  |  |
| Immunosuppression (vs. none) | 1.68 (1.02, 2.78) | 0.04 | 2.23 (1.23, 4.05) | 0.008 |
| **Admission investigations** |  |  |  |  |
| Haemoglobin <sup>(**)</sup> | 0.91 (0.86, 0.95) | <0.001 | 0.94 (0.88, 1.00) | 0.05 |
| Platelets <sup>(****)</sup> | 0.85 (0.75, 0.95) | 0.005 | 0.77 (0.67, 0.89) | <0.001 |
| Neutrophils <sup>(*)</sup> | 1.31 (1.17, 1.47) | <0.001 |  |  |
| Lymphocytes <sup>(+)</sup> | 0.58 (0.41, 0.83) | 0.001 |  |  |
| Sodium <sup>(**)</sup> | 1.14 (0.97, 1.35) | <0.001 |  |  |
| Potassium | 1.10 (0.92, 1.33) | 0.005 |  |  |
| Urea <sup>(+)</sup> | 12.3 (7.34, 20.5) | <0.001 | 2.67 (1.64, 4.32) | <0.001 |
| Creatinine <sup>(+)</sup> | 7.11 (3.75, 13.5) | <0.001 |  |  |
| C-Reactive Protein <sup>(***)</sup> | 1.19 (1.13, 1.25) | <0.001 | 1.15 (1.08, 1.22) | <0.001 |
| Alanine aminotransferase <sup>(+)</sup> | 0.59 (0.41, 0.86) | 0.006 |  |  |
| Alkaline phosphatase <sup>(+)</sup> | 2.63 (1.64, 4.21) | <0.001 | 2.53 (1.37, 4.67) | 0.003 |
| Bilirubin <sup>(+)</sup> | 1.31 (0.84, 2.04) | 0.23 |  |  |
| Albumin <sup>(*)</sup> | 0.67 (0.59, 0.76) | **<0.001** |  |  |
| Lactate |  |  | 2.67 (1.65, 4.33) | **<0.001** |
| Pulmonary infiltrates on chest radiograph (vs. none) | 1.27 (0.99, 1.64) | 0.07 | 1.89 (1.38, 2.60) | **<0.001** |
| **Admission bedside observations** |  |  |  |  |
| Heart rate <sup>(**)</sup> | 1.01 (0.96, 1.07) | **0.03** | 1.07 (1.00, 1.14) | 0.05 |
| Systolic BP <sup>(**)</sup> | 0.99 (0.94, 1.03) | **0.01** |  |  |
| Respiratory rate <sup>(**)</sup> | 1.45 (1.29, 1.63) | **<0.001** | 1.28 (1.08, 1.52) | **0.005** |
| Hypoxia (vs. none) | 1.77 (1.42, 2.20) | **<0.001** | 1.36 (1.03, 1.82) | **0.005** |
| Temperature | 0.93 (0.85, 1.03) | 0.17 |  |  |
| Glasgow Coma Score <15 (vs. 15) | 2.57 (1.98, 3.33) | **<0.001** | 1.92 (1.41, 2.62) | **<0.001** |

(\*) Hazard ratios given for a 5-unit increase in variable
(\*\*) Hazard ratios given for a 10-unit increase in variable
(\*\*\*) Hazard ratios given for a 50-unit increase in variable
(\*\*\*\*) Hazard ratios given for a 100-unit increase in variable
(+) Variable analysed on the log scale (base 10)
Variables found to be significant in univariable analysis but did not retain significance in multivariable analysis
were excluded from the final model
ACEi/ARB: angiotensin converting enzyme inhibitor/angiotensin II receptor blockers.

Age was found to be associated with the risk of death, with a 53% higher chance for each 10-year increase in age (adjusted hazard ratio (aHR) 1.53 [1.37–1.71]). No association was found with either sex or ethnicity. The absence of association between ethnicity and death persisted when Black, Asian and minority ethnic (BAME) groups were combined (see supplementary materials).

Respiratory disease (aHR 1.37 [1.03–1.81]) was associated with a higher risk of death as was immunosuppression (aHR 2.23 [1.23–4.05]). Conversely, no association was found with DM, hypertension, IHD, HF or use of ACEi, ARBs, or metformin.

Of the initial bedside observations, respiratory rate, hypoxia and low GCS retained a statistically significant association with death in multivariable analysis. Each increase in respiratory rate by 10 breaths/minute was associated with a 28% increase risk of death (aHR 1.28 [1.08–1.52]) and GCS <15 had an 92% increased risk of death (aHR 1.92 [1.41–2.62]). Hypoxia conferred a 36% increased risk of death (aHR 1.36 [1.03–1.82]).

Higher urea levels were associated with an increased risk of death (aHR 2.67 [1.64–4.32]), as were higher ALP (aHR 2.53 [1.37–4.67]), CRP (aHR 1.15 [1.08–1.22]) and lactate (aHR 2.67 [1.65–4.33]). Higher platelet counts were associated with a reduced risk of death (aHR 0.77 [0.67–0.89]).

The presence of pulmonary infiltrates on chest radiograph was associated with an increased risk of death (aHR 1.89 [1.38–2.60]).

In order to illustrate the difference in outcome between patients with different characteristics, patients were categorized into three tertiles based on the multivariable model. Survival at 60 days was 90% for the patients in the tertile with the ‘best’ combination of risk factors compared to 33% for the patients in the tertile with the ‘worst’ combination (Figure 3).

**Figure 3:**
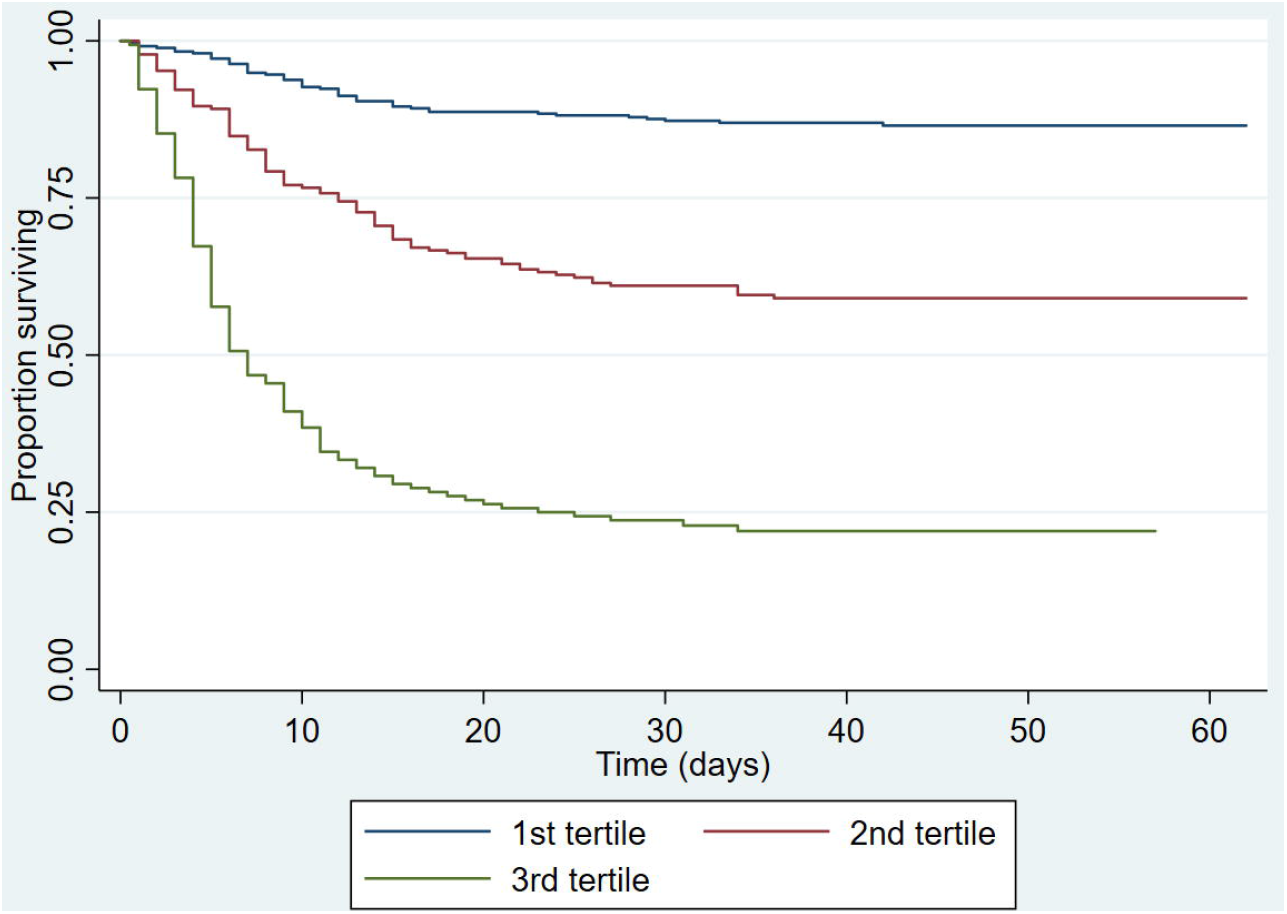
Kaplan-Meier plot of time to death split into tertiles by predicted risk from the multivariable model

A sensitivity analysis was performed excluding patients with a first positive swab >5days after admission. The same variables were included as in the original model and yielded comparable results (see supplementary material).

### Secondary Outcomes

Time to death or IMV was examined as a secondary composite endpoint using the same variables as for the primary outcome. Of the 981 patients in the cohort, 426 died or required IMV during the study period. The variables associated with this secondary outcome were similar to those for the primary endpoint, although ALP, respiratory disease and immunosuppressive medication were not found to be significantly associated. In addition, lower sodium and albumin levels were associated with higher risk of death or requiring IMV.

Four-hundred-and-sixty-one patients incurred the secondary endpoint of death, IMV or CPAP. The same variables were significantly associated with this endpoint as with the time to death or IMV, although the size of effects varied for some factors.

The full description of these secondary outcomes is shown in Table 3. Kaplan-Meier plots and sensitivity analyses are shown in the supplementary material.

**Table 3:**
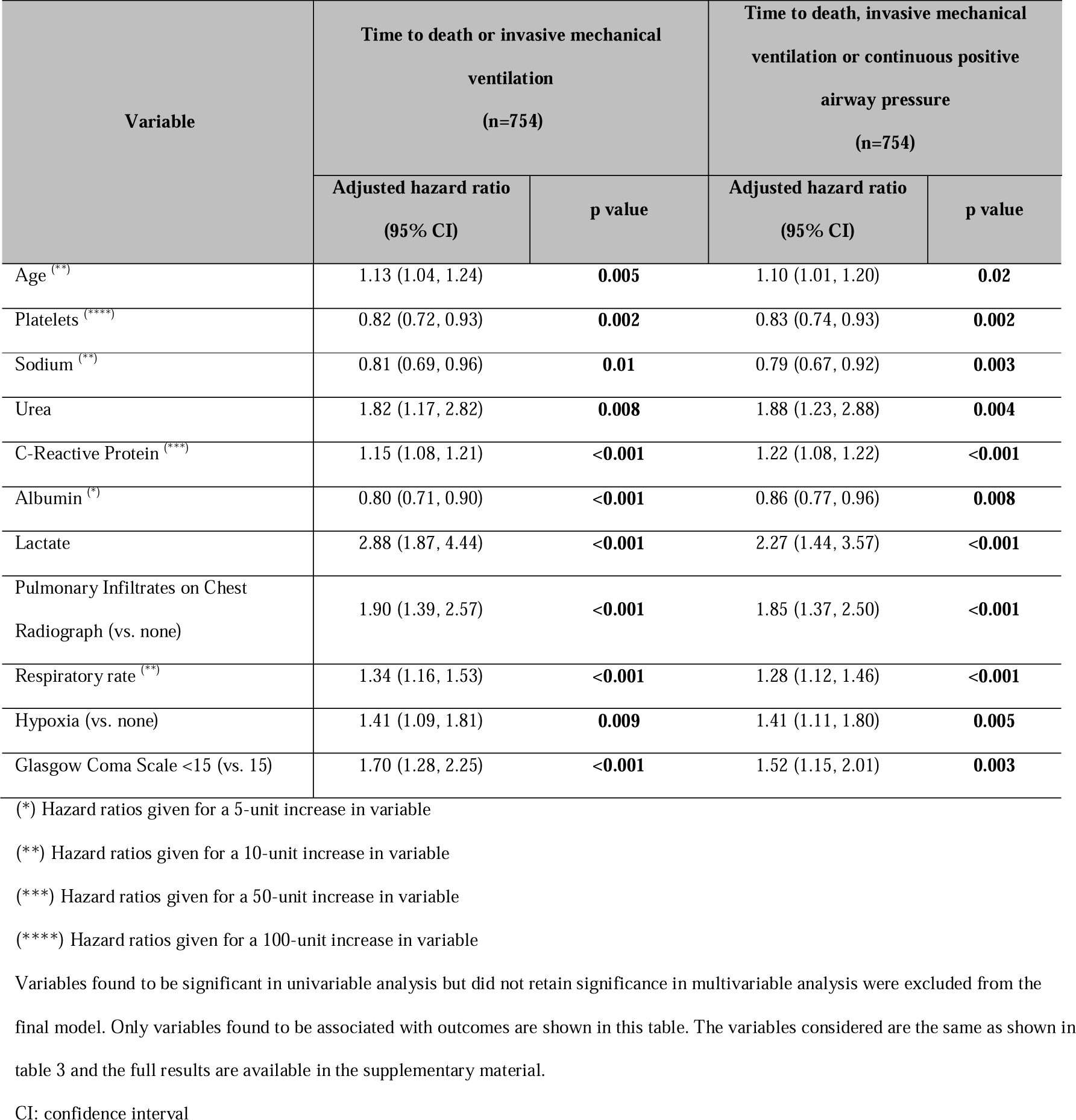
Secondary outcomes – multivariable analyses.

## Discussion

This large cohort study of patients hospitalized with COVID-19 describes the association between a number of clinical factors and the risk of severe disease. The study was conducted in a major London hospital that serves an ethnically diverse population, with 69.6% of study patients from Black, Asian and minority ethnic (BAME) groups. Overall, the mortality rates were comparable with those in other parts of the UK [14]. Although the results are from a single hospital, our catchment area covers a part of London with a very wide range of income, social status and ethnicity. We have no reason to believe that it is not representative of the city generally.

This study demonstrates a dramatic difference in outcomes between those who are in the tertile with the best combination of risk factors (90% survival at 60 days) versus those in the tertile with the worse combination of risk factors (33% survival). This provides clear evidence that risk-stratification can be made early in hospital presentation.

The results of this study generally corroborate a number of previously published findings. However, some differences also exist. Increasing age was strongly associated with poorer outcomes, corroborating the findings of others [3,6–8,17,18]. In line with previous research [19,20] a strong association between respiratory diseases and death was found. An association was also found between the use of immunosuppression and death which has been reported separately [21].

This population had a high incidence of hypertension and although this was a risk factor for severe disease in univariable analysis, it was not associated in multivariable analysis. This implies that although hypertension itself it not a risk factor for severe disease, it is associated with conditions which are. This is in line with other studies [7–9].

Unlike the recent UK cohort reported by the OpenSAFELY Collaborative [13], this study did not find an association between ethnicity and worse outcomes. Although BAME groups have been noted to have poor outcomes from COVID-19 at a population level, the explanation of which is likely to be a complex interaction between socio-economic and medical factors, the outcomes between different ethnic groups in hospitalized cohorts has often been found to be similar [9,22–24] and this was the case in our study.

In this cohort, 64.3% were male, which is notably higher than the hospital’s catchment area (50%) [15]. In line with a number of cohort studies of hospitalized patients, we failed to demonstrate an association with sex when adjusting for other factors [6,9,25]. One large UK cohort did find sex to be a significant factor when examined at the population level [14]. It may be that BAME and male patients are prone to more severe disease and therefore more of them appear in hospital-based cohorts but once in hospital they do not have higher mortality. Further work is needed to understand this.

Seventy-five percent of this cohort had pulmonary infiltrates reported on their admission chest radiograph which were associated with poorer outcomes as corroborated by other studies [26–28].

Several of the laboratory investigations taken on admission were associated with worse outcomes including CRP, urea, and ALP. Although several studies have found an association with raised hepatic enzymes [29–31], an independent association between death and ALP has not previously been reported. ALT was not associated with outcomes, suggesting that hepatocellular injury is not the principal explanation for this. Notably, the relationship between outcomes and ALP has been demonstrated in other infectious diseases and this finding merits further research [32].

Unlike some previous studies, reduced lymphocyte counts were not found to be associated with death [12]. An association was found between raised lactate and adverse outcomes. Although few other studies have included this variable, it has been previously reported [23,33]. Hyperlactataemia may represent hypoperfusion in patients with a sepsis-like presentation or a high adrenergic state secondary to immune activation and it should be explored in future research.

The results of the analyses on secondary endpoints were broadly comparable with the results of the primary endpoints, although neither the use of immunosuppression nor comorbidity with respiratory disease was significantly associated with the secondary outcomes. The dilution of this association may be explained by selection bias from clinicians on the suitability for either mechanical ventilation or CPAP for patients with these comorbidities.

This study has a number of strengths. Crucially, it included all sequentially admitted patients during a predetermined time period thus reducing the selection bias which has been seen in many studies. This study includes admission observations and investigations, accounting for disease severity in the model. It also benefits from a high degree of data completeness, with patient characteristics cross-validated with discharge summaries and other information sources to ensure accuracy.

A limitation of this study is that for the purpose of the time to event analyses, it had to be assumed that those who had clinically improved and were discharged prior to censoring remained alive until the censoring date. Although it is possible that some of these patients died post-discharge, this risk was minimized by ensuring that patients were screened for readmission and by ensuring that none of the cohort were discharged for community end-of-life care.

A second limitation is that a significant minority of patients (157/981) did not have an ethnicity documented. The majority of these patients had opted not to have their ethnicity recorded.

In conclusion, this large cohort study included all patients who were hospitalized with COVID-19 during the study period. It was conducted in an area of London with high levels of ethnic diversity [15]. When considering the high mortality among those hospitalized with COVID-19, early identification of those most susceptible to a severe disease is of paramount importance. This study found that predominantly respiratory features such as respiratory rate, oxygen saturations, infiltrates on chest radiograph and a history of respiratory disease were associated with severe disease. Comparatively few non-respiratory features were also identified as being associated with severe disease (age, reduced GCS and immunosuppression) along with a number of biochemical markers. For this new disease, understanding at the point of admission which patients are more likely to have a severe disease course is key to making appropriate and timely treatment escalation decisions, and ultimately reducing avoidable mortality.

## Data Availability

STATA codes can be obtained from the authors. Data on all COVID-19 admission has already been stared with Public Health England. Application for further patient level data should be addressed to Northwick Park Hospital's research ethics committee.

## Author Contributions

NV, DLC, and TANR designed the study. MA, JG, TANR collected the data with assistance from those acknowledged below. PB, and JG conducted the data analysis. JG wrote the first draft of the article and conducted the literature search. TANR, MA, PB, AMW, DLC and NV all reviewed and approved the final report. NV and DLC are joint senior authors.

## Acknowledgements

We thank Dr Poppy Utting, Dr Megan Miscampbell, Dr Dorina Condurache, Dr Nathan Eager, and Dr Krisha Shah for their help in collecting patient data. We are grateful to all our colleagues at London North West Hospitals University Healthcare NHS Trust for their outstanding commitment to maintaining good clinical care during the COVID-19 pandemic and for their support of this project.

